# segcsvd_WMH_: A convolutional neural network-based tool for quantifying white matter hyperintensities in heterogeneous patient cohorts

**DOI:** 10.1101/2024.06.20.24309230

**Authors:** Erin Gibson, Joel Ramirez, Lauren Abby Woods, Julie Ottoy, Stephanie Berberian, Christopher J.M. Scott, Vanessa Yhap, Fuqiang Gao, Roberto Duarte Coello, Maria Valdes Hernandez, Anthony E. Lang, Carmela M. Tartaglia, Sanjeev Kumar, Malcolm A. Binns, Robert Bartha, Sean Symons, Richard H. Swartz, Mario Masellis, Navneet Singh, Alan Moody, Bradley J. MacIntosh, Joanna M. Wardlaw, Sandra E. Black, ONDRI Investigators, ADNI, CAIN Investigators, colleagues from the Foundation Leducq Transatlantic Network of Excellence, Andrew SP Lim, Maged Goubran

## Abstract

White matter hyperintensities (WMH) of presumed vascular origin are an MRI-based biomarker of cerebral small vessel disease (CSVD). WMH are associated with accelerated cognitive decline and increased risk of stroke and dementia, and are commonly observed in aging, vascular cognitive impairment, Alzheimer’s and Parkinson’s disease, and related dementias. The accurate, reliable, and rapid measurement of WMH in large-scale multi-site clinical studies with heterogeneous patient populations remains challenging. The diversity of MRI protocols and image characteristics across different studies as well as the diverse nature of WMH, in terms of their highly variable shape, size, distribution, and underlying pathology, adds additional complexity to this task. Here, we present segcsvd_WMH_, a novel convolutional neural network-based tool for quantifying WMH. segcsvd_WMH_ is specifically designed for accurate and robust performance when applied to diverse clinical patient datasets. Central to the development of this tool is the curation of a large patient dataset (>700 scans) sourced from seven multi-site studies, encompassing a wide range of clinical populations, WMH burden, and imaging parameters. The performance of segcsvd_WMH_ is evaluated against three widely used WMH segmentation tools, where we demonstrate significantly enhanced accuracy and robustness across a range of challenging conditions and datasets.

## Introduction

White matter hyperintensities (WMH) of presumed vascular origin, are commonly observed on brain magnetic resonance imaging (MRI) in older adults and patients with neurovascular and neurodegenerative disease (**Wardlaw et al., 2013**). Visualized as hyperintense (bright) signals on T2-weighted Fluid-Attenuated Inversion Recovery (FLAIR) MRI scans, WMH are a key imaging biomarker of cerebral small vessel disease (CSVD) and are associated with a range of poor clinical outcomes, including accelerated cognitive decline and increased risk for stroke and dementia (**Prins and Scheltens, 2015**). Accurate detection and quantification of WMH are crucial for diagnosis, monitoring disease progression, and research into the underlying mechanisms of these conditions (**Wardlaw et al., 2019)**, necessitating automated, reliable, and efficient tools for measurement of WMH.

Recent advances in machine learning, particularly in the field of deep learning and specifically convolutional neural networks (CNN) (**Derry et al., 2023**), have greatly expanded the availability of automated WMH segmentation tools. Despite these advancements, significant challenges remain in their effective application across large and diverse clinical datasets.

One challenge lies in the heterogeneity of MRI data across diverse clinical and research datasets. Factors such as image resolution, contrast properties, and the presence of artifacts can substantially influence the accuracy of WMH detection (**De Guio et al., 2016**). Furthermore, the diverse nature of WMH, in terms of their highly variable shape, size, distribution, and underlying pathology further complicates the segmentation task.

These challenges often manifest as inconsistent segmentation performance across different datasets. A common issue is poor generalization, characterized by a decline in performance when existing tools are applied to new datasets with different imaging or patient characteristics (**Wang et al., 2023**). To achieve comparable performance levels across datasets, these tools often require semi-automated parameter tuning, such as adjustments to the threshold that converts model probabilities into binary segmentations. In some cases, the model itself must be fine-tuned or re-trained to adapt to the unique characteristics of a new dataset. These dataset-specific adjustments require considerable time and resources and are not always feasible or successful.

Given the challenges involved in WMH segmentation, there is a continued demand for tools that provide greater accuracy and robustness across diverse imaging datasets. This work aims to address this demand through the development of segcsvd_WMH_, a novel WMH segmentation tool designed specifically for enhanced segmentation performance in heterogeneous clinical datasets characterized by highly varied imaging parameters, patient types, and WMH burden.

Central to the development of segcsvd_WMH_ was the curation of a large patient dataset (>700 scans) sourced from seven multi-site studies, encompassing a range of clinical populations, WMH burden, and imaging parameters. This dataset was further enriched by the creation of highly accurate ground truth labels. Initially synthesized from two legacy WMH segmentation tools (**Gibson et al., 2010; Mojiri Forooshani et al., 2022**), these labels were meticulously refined through a semi-automated procedure consisting of precise manual thresholding and/or editing.

segcsvd_WMH_ features an innovative two-stage approach to WMH segmentation. In the first stage, SynthSeg, a recently validated CNN-based tool (**Billot et al., 2023**) from the FreeSurfer software suite (**Fischl, 2012**) is utilized. This tool was developed using a large synthetic dataset and provides fast and robust segmentation of non-WMH cortical and subcortical structures for diverse clinical scans of any contrast and resolution. segcsvd_WMH_ leverages the SynthSeg segmentation to generate a regional mask consisting of hippocampal and sulcal cerebrospinal (sCSF) voxels. This regional mask provides valuable spatial context to inform and constrain the subsequent WMH segmentation task, where the hippocampus acts as a key anatomical reference point and sCSF serves as a marker for the perimeter of cortical gray matter. In the second stage, the regional mask and FLAIR image are used as dual inputs to a CNN optimized for the task of WMH segmentation. Minimal preprocessing, consisting of brain masking, bias correction, and intensity standardization, is performed to enhance consistency across datasets prior to inputting the FLAIR images into the CNN. This two-stage approach explicitly combines relevant anatomical information with the contrast characteristics of the FLAIR image to improve the accuracy and sensitivity of the WMH segmentation.

Several advanced training strategies were integrated into the development of segcsvd_WMH_ to ensure robust WMH segmentation. These include the creation of three distinct models, each fine-tuned by adjustment of Tversky loss function parameters to achieve unique precision-sensitivity weightings. This fine-tuning yielded a diverse collection of models, each with varied yet capable performance characteristics. In addition, segcsvd_WMH_ employs sophisticated data augmentation strategies, both during training and prediction, to further enhance its segmentation capabilities across datasets.

The performance of segcsvd_WMH_ was evaluated by benchmarking it against three previously validated segmentation tools: HyperMapp3r (**Mojiri Forooshani et al., 2022**), SAMSEG (**Puonti et al., 2016**), and WMH-SynthSeg (**Laso et al., 2024**). These tools were all developed with the aim of achieving robust performance on diverse clinical datasets without retraining, and are easily accessible, either distributed within the FreeSurfer suite or available for direct download, providing ideal benchmarks in this context. In addition to benchmarking against these tools, other key aspects of segmentation performance were evaluated, including consistency across different binary segmentation thresholds, robustness to simulated MR artifacts, and adaptability to diverse and unseen data. Collectively, these evaluations provided a comprehensive assessment of segcsvd_WMH_ segmentation performance across a variety of challenging datasets and conditions.

## Results

This section begins with a detailed description of the datasets used to develop and validate segcsvd_WMH_ and continues with the calculation of WMH volumes and FLAIR WMH contrast ratios to quantify the degree of variability in WMH burden and FLAIR contrast properties across the included datasets. Subsequent sections focus on evaluating the performance of segcsvd_WMH_ against the three benchmark tools using several metrics. These include an overlap metric (Dice score) which evaluates the agreement with ground truth voxels; a boundary-based metric (normalized surface distance) which assesses accuracy in the shape of the segmented objects; a volume-based metric (average volume difference) which quantifies the accuracy of volume estimation; and two additional metrics (sensitivity and precision) which assess accuracy based on the proportion of false negative or false positive voxels.

Collectively, these metrics provide a comprehensive assessment of segmentation accuracy at the pixel, shape, and volume level, in accordance with established recommendations for the evaluation of pixel-level segmentation tasks (**Maier-Hein et al., 2024**).

### Description of datasets

A large dataset consisting of 947 3T FLAIR images and FLAIR-based SynthSeg segmentations was assembled from seven different studies (**Table 1**). Many of these studies were large multi-site projects, and two (ADNI/LD n=85/70) employed newer 3D FLAIR imaging protocols with 1 mm isotropic, or approximately isotropic, voxel resolution. These studies spanned a wide range of patient populations, including individuals with Alzheimer’s disease, vascular cognitive impairment, aphasia, sleep apnea, carotid stenosis, as well as normal controls. Altogether, this dataset consisted of a highly heterogeneous set of images, encompassing a wide range of imaging parameters and patient populations.

**Table 1.**
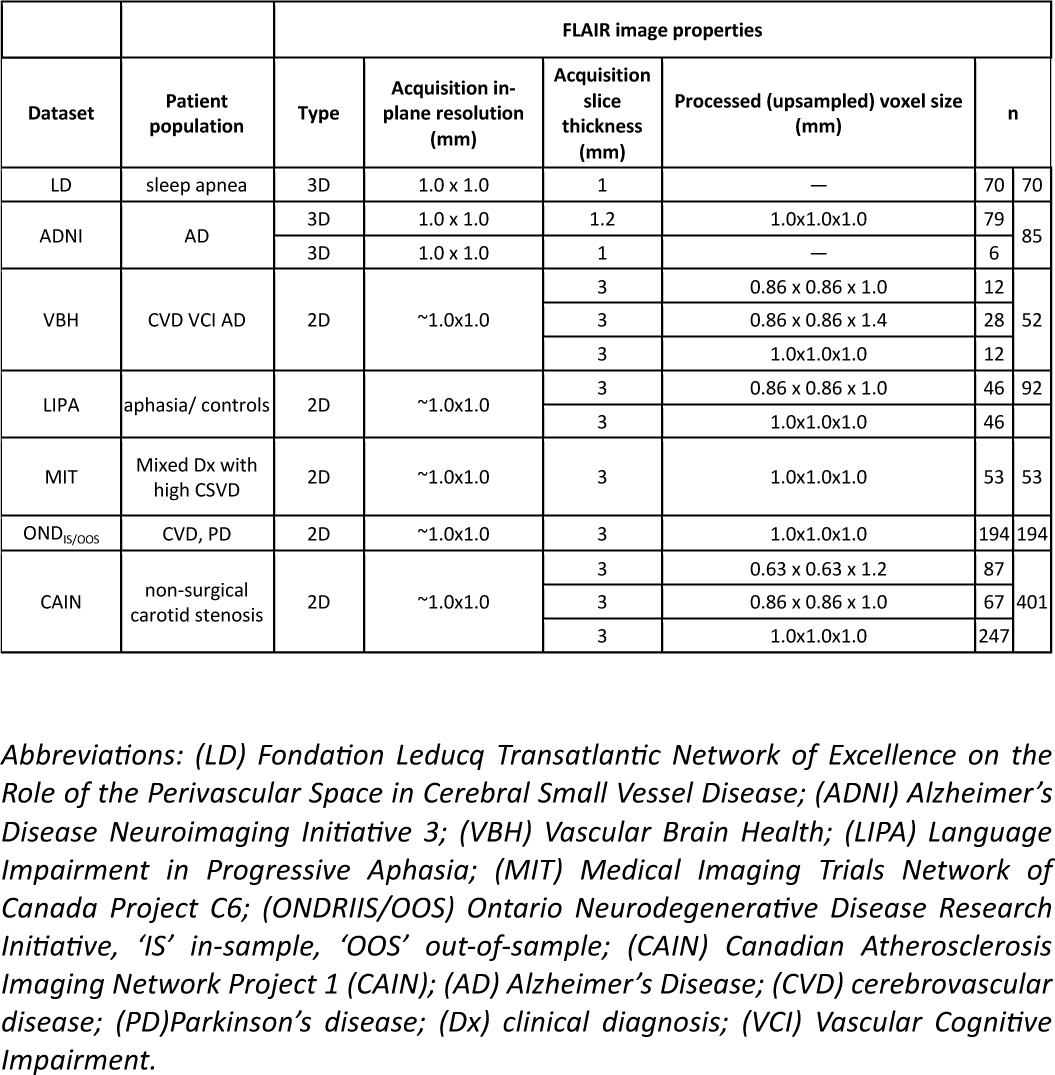
Key imaging characteristics for each dataset, including the resolution of the acquired and processed FLAIR data, a critical factor influencing model performance. Other MR acquisitions parameters that affect FLAIR contrast are also important factors, and these varied by study and site. To provide a more direct and interpretable assessment of these contrast differences, FLAIR WMH contrast ratios were calculated for each dataset and are reported in the results section.

The full dataset (n=947) was partitioned into separate training (n=781), validation (n=12), and test (n=154) datasets. A minimal amount of data was allocated to the validation dataset, as it was used only to select the binary segmentation threshold and to set coarse limits for the random selection of models for ensembling. For model validation purposes, the test data were further subdivided into four separate datasets based on two criteria: whether they were included (“in-sample”) or excluded (“out-of-sample”) from the training dataset, and whether they were acquired with approximately isotropic 1mm voxels (3D) or with 3mm thick slices (2D). This resulted in four separate test datasets: 2D_IS_ (2D in-sample; n=18); 2D_OOS_ (2D out-of-sample; n=41); 3D_IS_ (3D in-sample; n=10); and 3D_OOS_ (3D out-of-sample; n=5 with and n=80 without ground truth segmentation), where the in-sample datasets provided a test of performance on familiar data, and the out-of-sample datasets provided a test of generalization performance on unseen data.

A second set of test datasets was generated by applying a transform to each image (FLAIR and FLAIR-based SynthSeg segmentation) in the first set of test datasets. This transform involved swapping the x and z axes, and then updating the orientation field in the image header to incorrectly reflect this change. Specifically, all images were in LPI or RPI format initially, and were transformed to IPL or IPR format, with the orientation field in the image header updated to indicate SPL or SPR format. This produced a set of images formatted in an orientation that was neither present in the training data nor discernible from the image header description, allowing for a rigorous test of whether model performance was entirely independent of orientation.

A third test dataset was created to augment the 3D_OOS_ test dataset and assess segmentation performance in the presence of simulated MR artifacts. This was accomplished by randomly introducing five levels of spike noise artifact to each FLAIR image, resulting in a total of 30 FLAIR images for this dataset, each with ground truth segmentations. Following the addition of spike noise, the SynthSeg segmentations were regenerated, thereby exposing all components of the segcsvd_WMH_ tool to the effects of simulated spike noise.

### Variation in WMH burden and contrast ratios across datasets

The degree of WMH burden across datasets was evaluated by expressing the ground truth WMH volumes as a percentage of the total intracranial volume (**Figure 1A**). A high degree of variability in these volumes was observed both across and within datasets, ranging from 0.1% to 7% of total intracranial volume, indicating substantial heterogeneity in CSVD severity across individuals and datasets included in this work.

**Figure 1.**
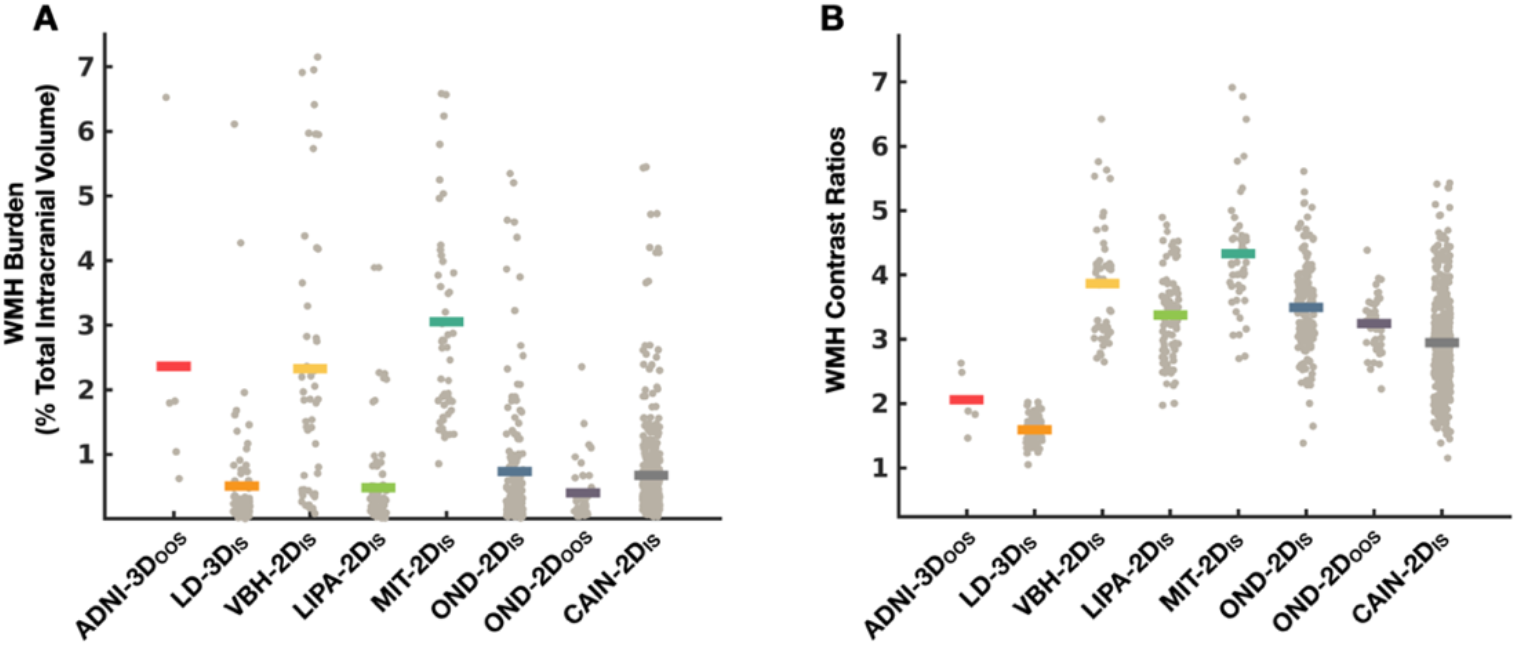
Variation in WMH burden (A) and WMH contrast ratios (B) across datasets. Mean values are represented by solid lines and individual data points are plotted in grey. A high degree of variation was present in both measures, indicating substantial heterogeneity in the data used to develop and validate segcsvd_WMH_..

Differences in FLAIR contrast properties across datasets were assessed using WMH contrast ratios, which quantified the visibility of WMH relative to gray matter (GM) and white matter (WM) (**Figure 1B**). The LD 3D_IS_ and ADNI 3D_OOS_ datasets, which employed isotropic or nearly isotropic FLAIR imaging protocols, exhibited significantly lower WMH contrast ratios than all other datasets (permuted p-values < 0.05), indicating reduced visibility of WMH on FLAIR for these datasets

### Segmentation performance for the standard test data

#### Agreement with ground truth segmentations

Overall, segcsvd_WMH_ was the only tool that maintained a consistently high level of performance across all metrics on the standard test data (**Figure 2; first column**). To facilitate a more direct performance comparison, the mean difference for each performance metric was calculated between segcsvd_WMH_ and the benchmark tools for the standard test data (**Table 2**). Permutation tests revealed that segcsvd_WMH_ significantly outperformed the benchmark tools in almost all instances. Often these performance improvements were quite substantial. For example, segcsvd_WMH_ exhibited mean Dice score enhancements across test datasets of: 8.0%+-9.7% over HyperMapp3r; 23.0%+-7.6% over SAMSEG; and 45.0%+-5.5% over WMH-SynthSeg. Notably, there were only two instances where a benchmark tool significantly outperformed segcsvd_WMH_. HyperMapp3r exhibited significantly higher sensitivity on the 2D_OOS_ dataset, however this result was expected given that 38 of the 41 scans in that dataset were used to train HyperMapp3r, thereby inflating its sensitivity for familiar data. HyperMapp3r also exhibited significantly higher precision for the 3D_OOS_ dataset, but this was offset by worse performance for all other metrics, indicating that the enhanced precision for this dataset was associated with an overall decrease in performance.

**Table 2.**
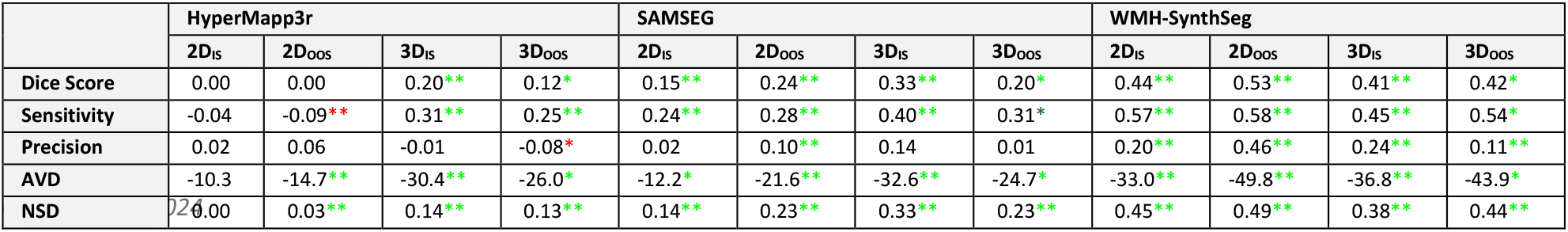
Mean performance differences between segcsvd_WMH_ and the three benchmark tools for the standard test data. Asterisks signify significant differences, with a single asterisk (*) indicating p < 0.05 and double asterisks (**) indicating p < 0.001. Asterisk color indicates the direction of the performance effect, with green signifying performance improvements, and red indicating performance declines, for segcsvd_WMH_ compared to the benchmark tools.

**Figure 2.**
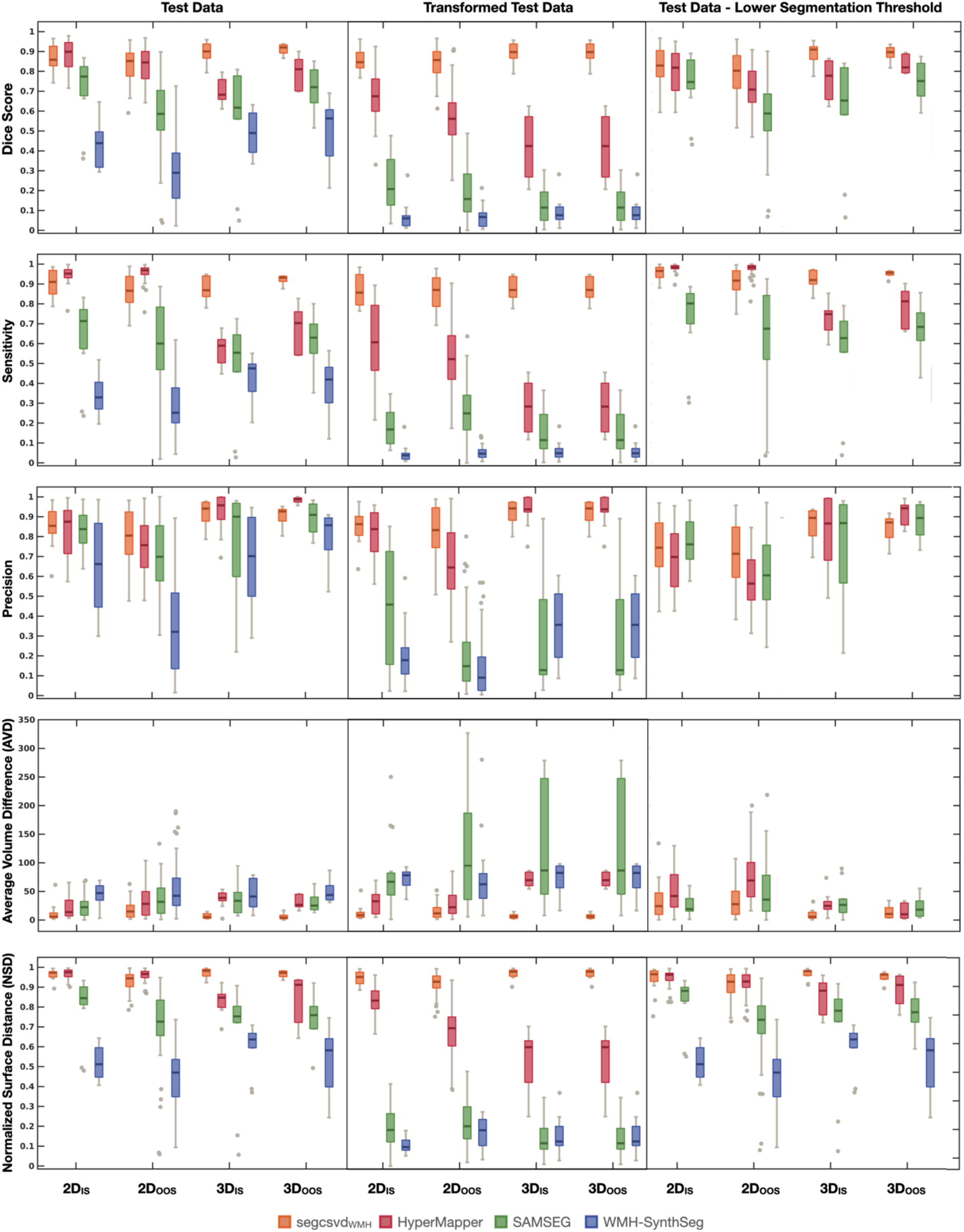
Performance across three test datasets for the four segmentation tools. The first column shows performance on the standard test data. The second column shows performance on the transformed test data after reorientation with inaccurate orientation information embedded into the image header. The last column shows performance on the standard test data using a lower binary segmentation threshold of 0.1. For clarity and ease of comparison, the AVD plots were truncated from 750 to 350 and do not display all outlier points for SAMSEG and WMH-SynthSeg. segcsvd_WMH_ was the only tool to maintain a consistently high level of performance across all metrics and test datasets.

#### Agreement with ground truth volumes

The agreement with ground truth volumes was analyzed for each tool in several regions of interest (ROIs) that included either periventricular, deep, or total WMH voxels (**Figure 3**). segcsvd_WMH_ was the only tool that exhibited strong and stable correlations with ground truth volumes across all ROIs (mean r=0.99+-0.007). In contrast, the benchmark tools tended to exhibit either slightly weaker correlations (e.g. SAMSEG for all ROIs; mean r=0.93+-0.05) that were biased toward underestimation of the true volumes, or much weaker correlations (e.g. HyperMapp3r for deep WMH; mean r=0.53+-0.18 and WMH-SynthSeg for all ROIs; r=0.74+-0.08). In general, the benchmark tools were more prone to underestimation than segcsvd_WMH_, and this tendency was also clearly apparent upon visualization of their segmentation outputs (**Figure 4**).

**Figure 3.**
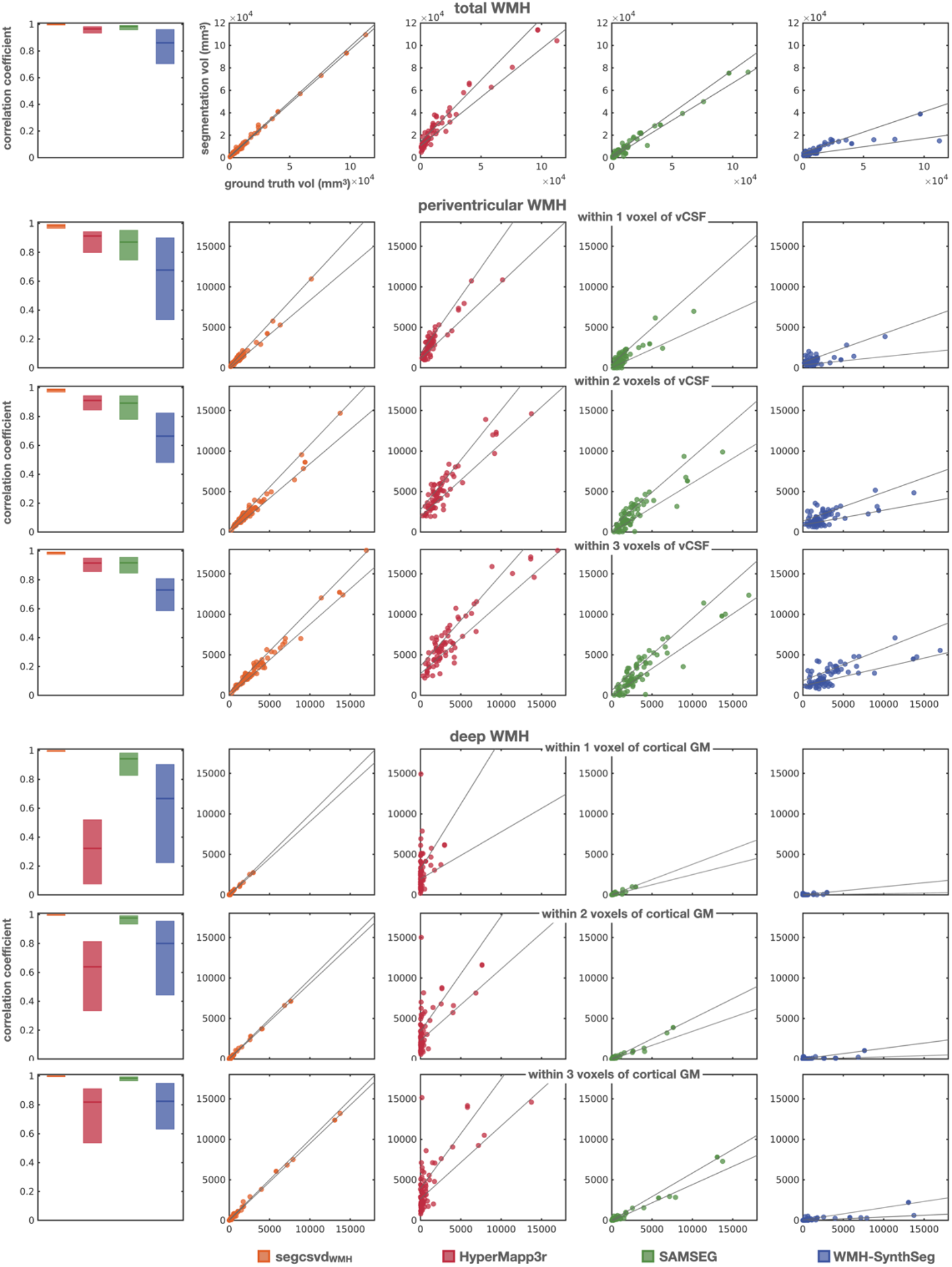
Relationship with ground truth volumes for total, periventricular and deep WMH. First column displays the Pearson correlation coefficients (solid line) with their 95% bootstrap estimated confidence intervals (shaded area). Remaining columns display the corresponding scatterplots for these correlations with their 95% bootstrap estimated confidence intervals (solid lines), representing the upper and lower bounds of the regression line. Scatterplots include data from all test datasets with available ground truth segmentations (n=74). segcsvd_WMH_ was the only tool that exhibited strong, stable, and unbiased correlations with ground truth volumes consistently across all ROIs. In contrast, the benchmark tools tended to exhibit lower and more variable performance across ROIs. This was particularly evident for HyperMapp3r, which showed good performance for global and periventricular WMH, but substantially lower performance for deep WMH.

**Figure 4.**
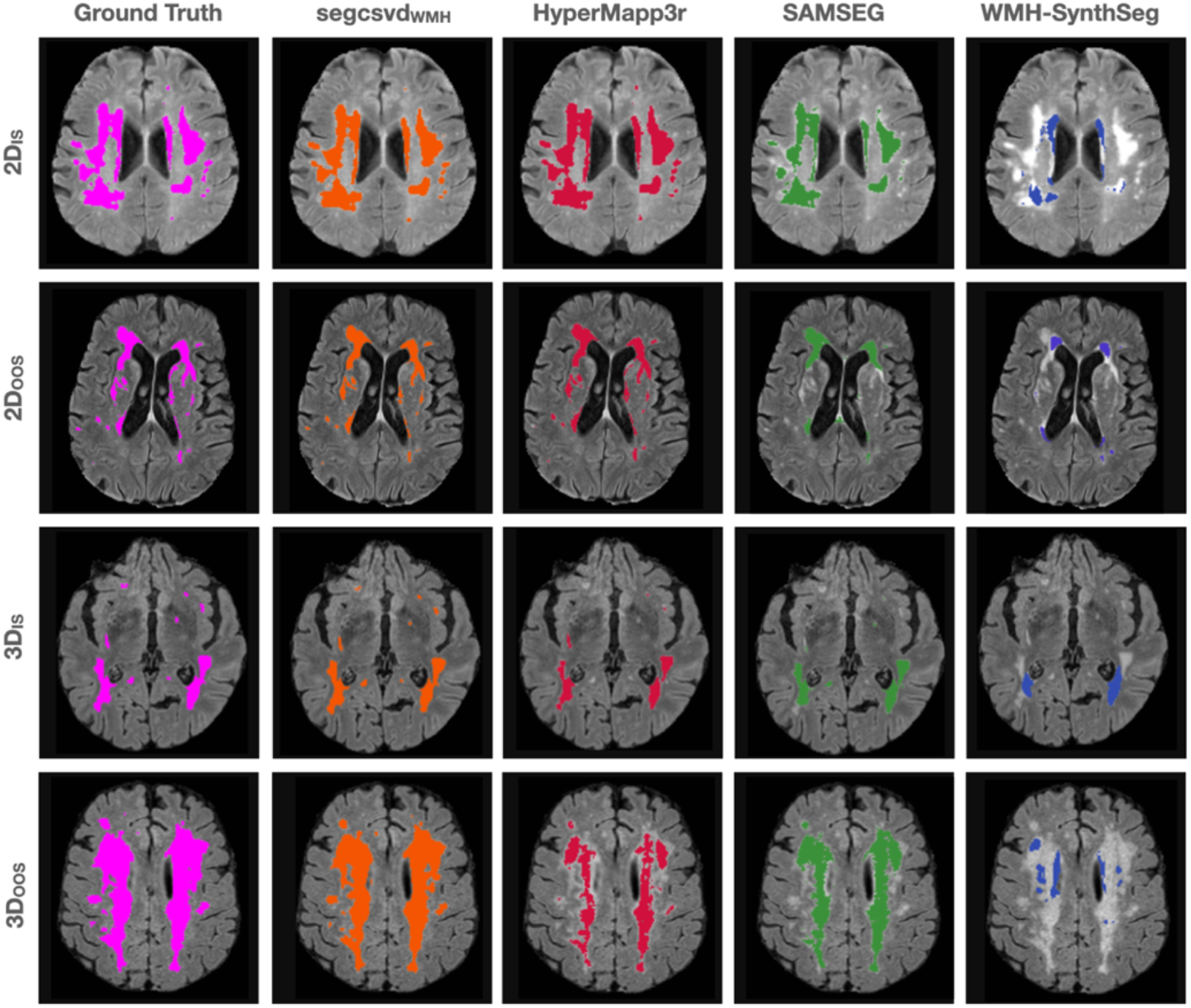
Illustrative examples of the segmentation output for each tool. segcsvd_WMH_ demonstrated robust performance on the test data, showcasing consistently high agreement with ground truth segmentations and less underestimation than the benchmark tools.

A follow-up analysis was performed on the total WMH volumes, stratified into three levels based on the ground truth segmentation volumes, corresponding to low, moderate, or high WMH burden. Correlations with ground truth volumes across these three levels were stronger and more consistent for segcsvd_WMH_ (mean r=0.93+-0.07) compared to HyperMapp3r (mean r=0.68+-0.28), SAMSEG (mean r=0.72+0.22), and WMH-SynthSeg (mean r=0.51+-0.24). Thus, segcsvd_WMH_ exhibited greater agreement with total WMH volumes, both across all levels, and at each level, of WMH burden, indicating strong and consistent performance across all WMH severity levels.

### Segmentation performance for the transformed test data

Performance metrics were calculated for the transformed test data, after reorientation with inaccurate orientation information embedded into the image header (**Figure 2; second column**). For segcsvd_WMH_, performance on the transformed data was in line with performance on the standard test data across all metrics and test sets (**Figure 2; first column**). In contrast, for the benchmark tools, there was a marked decline in performance on the transformed data across metrics and test sets

### Segmentation performance for standard test data with lower segmentation threshold

For segcsvd_WMH_, HyperMapp3r and SAMSEG, a secondary analysis was performed using a lower binary segmentation threshold of 0.1 for the standard test data to determine whether any observed WMH underestimation for the benchmark tools could be improved using a more inclusive threshold (**Figure 2; last column**). This yielded a similar overall pattern of results as with the original (default) thresholds (**Figure 2; first column**), indicating that the lower sensitivity of the benchmark tools was not simply due to the use of an overly conservative threshold.

### Robustness to different binary segmentation thresholds

Performance metrics were evaluated at 20 different binary segmentation thresholds for both segcsvd_WMH_ and HyperMapp3r (**Figure 5**). segcsvd_WMH_ demonstrated high stability in its performance metrics. For example, the Dice score for segcsvd_WMH_ remained consistently high at thresholds of 0.1 or higher across all test datasets. Importantly, for any given threshold, the performance metrics for segcsvd_WMH_ showed a high degree of uniformity across test datasets, indicating robust and consistent segmentation outcomes independent of the threshold. In contrast, the optimal binary segmentation threshold was highly dependent upon the dataset for HyperMapp3r (e.g. max Dice Score of 0.87 at a threshold of 0.7 for 2D_OOS_, versus max Dice Score of 0.83 at a threshold of 0.05, decreasing to 0.65 at a threshold of 0.7, for 3D_OOS_).

**Figure 5.**
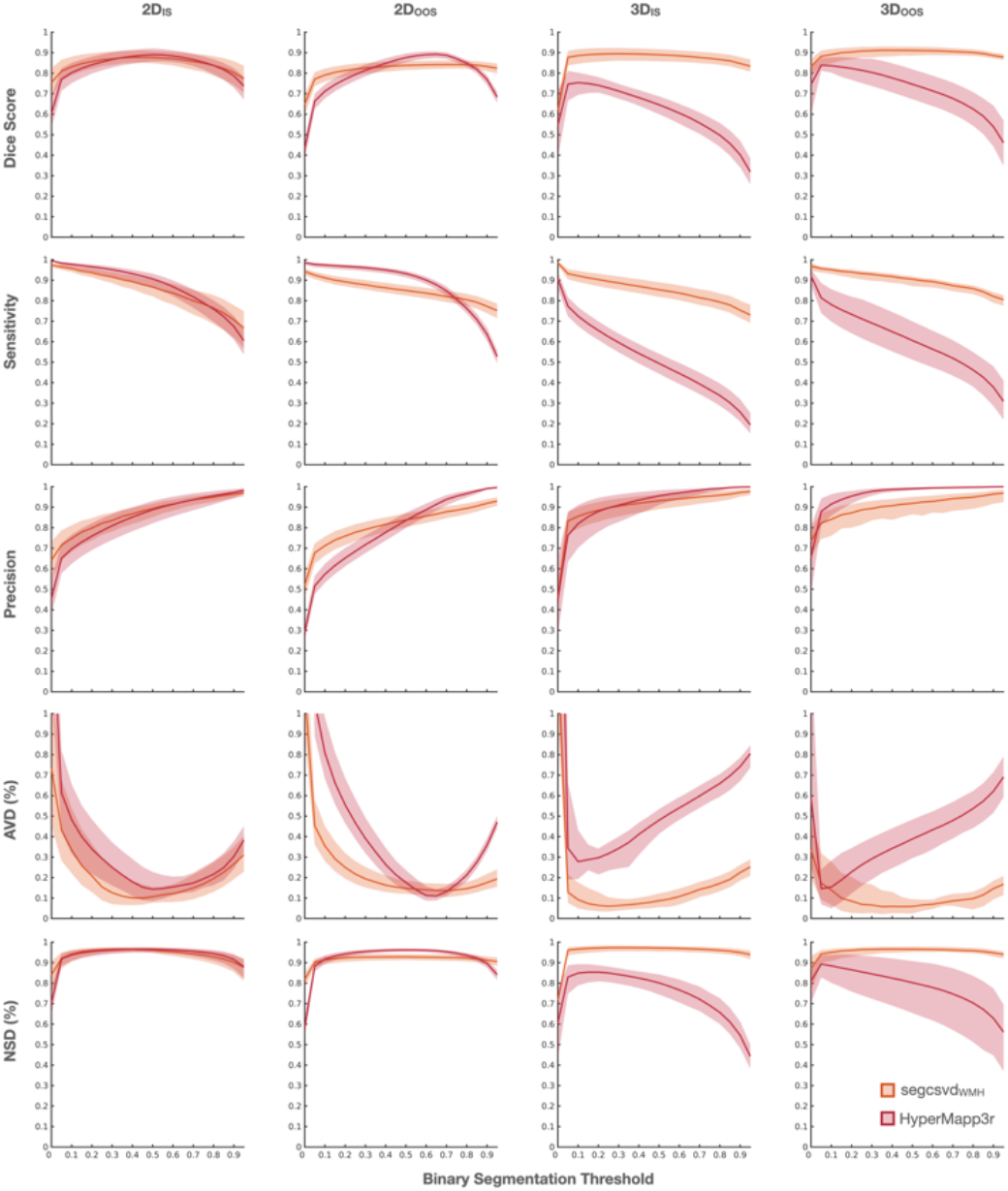
Performance comparison for segcsvd_WMH_ and HyperMapper across 20 binary segmentation thresholds. Mean performance metrics are plotted with shaded areas signifying the 95% bootstrap estimated confidence intervals. Performance metrics for segcsvd_WMH_ (orange) exhibited much greater stability in response to changes in the binary segmentation threshold than HyperMapper (red), and this stability was largely maintained across the test datasets.

### Qualitative assessment of segmentation performance for the 3D_OOS_ dataset

For each scan in the 3D_OOS_ ADNI test dataset without ground truth labels (n=80), the segmentation output for the three top-performing tools was anonymized and visually ranked in terms or their relative accuracy. segcsvd_WMH_ ranked as most accurate for 100% of the scans. Conversely SAMSEG and HyperMapp3r ranked as least accurate, for 91.8% and 8.2% of the scans, respectively.

### Robustness to simulated MRI artifacts

The normalized Mattes mutual information (NMMI) metric was used to quantify the severity of the simulated spike noise artifact (**Figure 6)** and the performance of each tool at six levels of artifact severity was assessed (**Figure 7)**. Overall, segcsvd_WMH_ exhibited highly robust performance in the presence of mild to moderate spike noise artifacts, with minimal change in its performance metrics between severity levels 0 (no artifact) and 3 (moderate artifacts). Its performance decreased substantially at the higher severity levels 4 and 5. In contrast, the performance of SAMSEG progressively decreased as the severity increased, while HyperMapp3r and WMH-SynthSeg exhibited the greatest robustness to simulated spike noise artifacts, maintaining higher consistency in their performance metrics across all levels, however, the overall performance of WMH-SynthSeg was consistently low. Consequently, in the context of simulated spike noise, HyperMapp3r emerged as most effective for data with severe artifacts (levels 4 and 5), while segcsvd_WMH_ proved most effective for data with minimal to moderate artifacts (levels 0 to 3).

**Figure 6.**
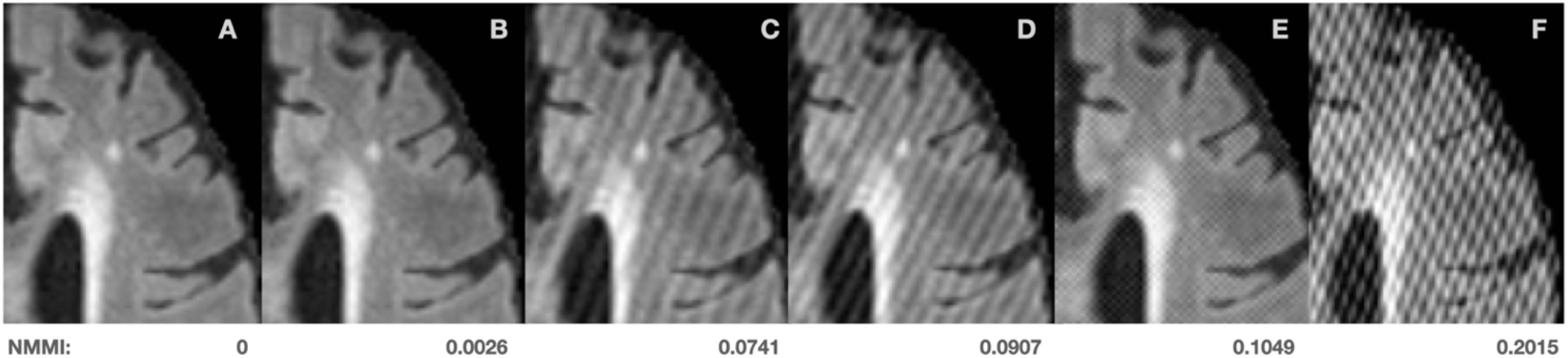
Illustrative example of the simulated spike noise artifacts added to the uncorrupted FLAIR data (A). The normalized Mattes mutual information (NMMI) metric served as an objective measure of artifact severity. This metric was implemented within a minimization framework where smaller values indicate greater similarity. In the current context, low NMMI values indicate minimal visible artifacts (B). Moderate NMMI values reflect visible artifacts, more typical of those often observed on actual MR scans (C,D). High NMMI values correspond to highly visible artifacts characterized by a pronounced cross-hatching pattern, less typical of those observed on actual MR scans (E,F).

**Figure 7.**
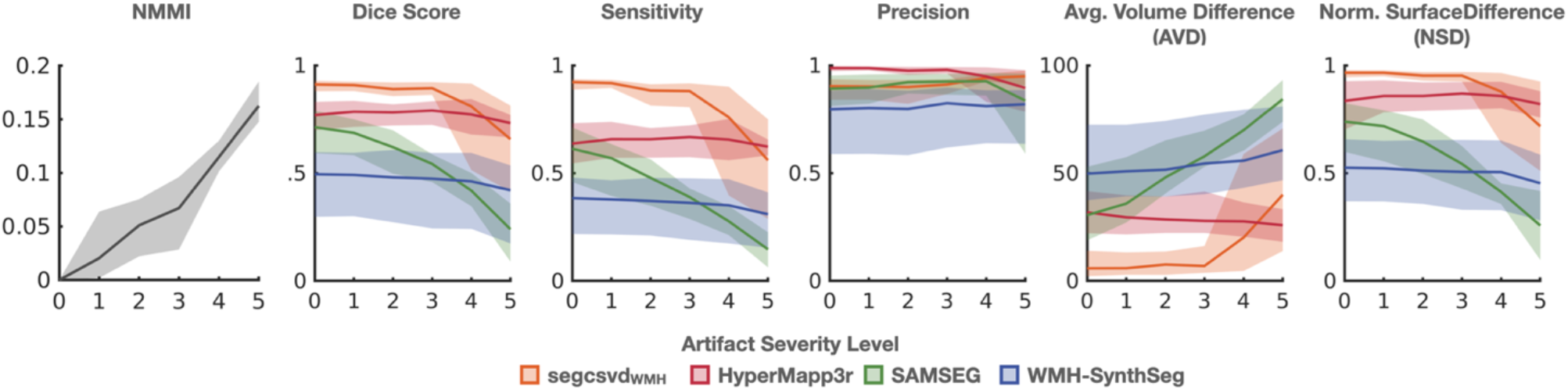
Performance of each tool across the six levels of simulated spike noise artifact. Artifact severity was quantified using the normalized Mattes Mutual Information metric (NMMI), stratified into 5 levels of increasing severity (plot 1). Mean values are represented by solid lines, with shaded areas indicating the 95% bootstrap estimated confidence intervals (plots 2-6). Overall, segcsvd_WMH_ exhibited the strongest and most stable performance for lower levels of artifact (severity levels 0 through 3). In contrast, HyperMapper exhibited the strongest and most stable performance for higher levels of artifact (severity levels 4 and 5).

Individual Dice scores for this test dataset were visualized to further examine the impact of artifact severity on segmentation performance (**Figure 8**). This highlighted the strong and stable performance of segcsvd_WMH_ at low to moderate levels of simulated artifact (NMMI < ∼0.125), but also revealed its potential for segmentation failures (Dice scores less than ∼0.6), at higher levels of simulated artifact (NNMI > ∼0.125). In contrast, while the Dice Scores for HyperMapp3r tended to be lower overall, no such segmentation failures were observed.

**Figure 8.**
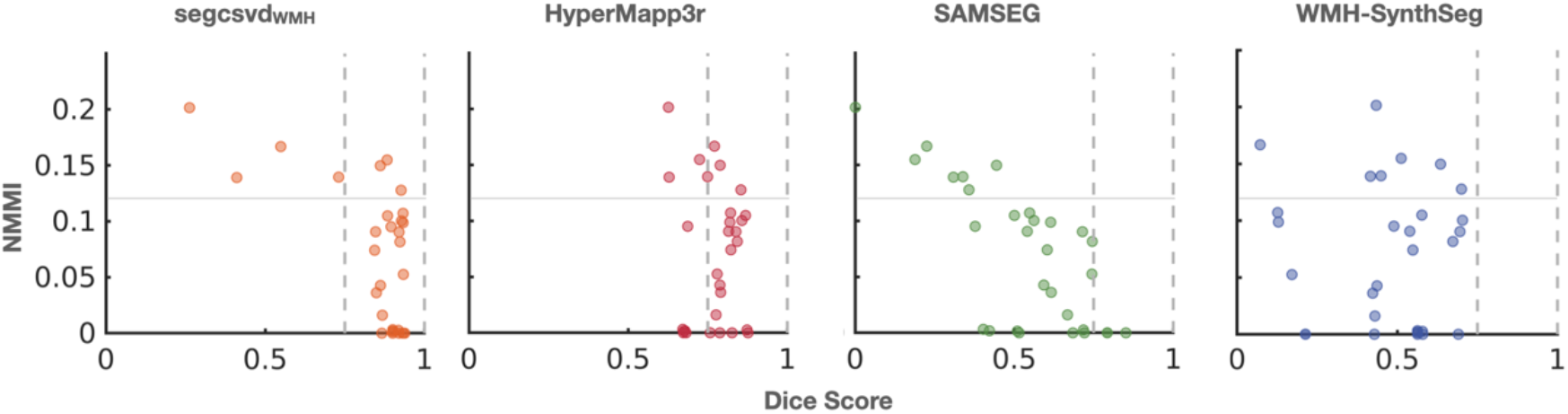
Dice scores for the test dataset with simulated spike noise artifacts. Dotted lines correspond to a Dice score of 0.8 and solid lines correspond to a NMMI of 0.125. Overall, segcsvdWMH exhibited strong and robust performance, with consistently high Dice Scores for low to moderate levels of simulated spike noise artifact (NMMI < ∼0.125). However, segmentation failures for segcsvdWMH were observed at higher levels of simulated spike noise artifact (NMMI >∼0.125), with Dice scores below ∼0.6.

## Discussion

This work introduced segcsvd_WMH_, an innovative CNN-based tool designed specifically for enhanced WMH segmentation on FLAIR images in heterogeneous clinical datasets with varying degrees of CSVD severity. This tool represents a significant improvement over existing segmentation methods, providing greater accuracy and consistency in performance across a wide range challenging conditions and datasets. These improvements, along with key innovations and contributions, are detailed in the following sections.

### High-fidelity ground truth dataset

Critical to the success of segcsvd_WMH_ was the creation of a large, diverse, and highly accurate ground truth dataset. This multi-study dataset, comprising over 700 individual FLAIR images, featured a wide range of CSVD severities from various patient populations and healthy controls, and diverse imaging protocols, including newer 3D isotropic FLAIR protocols characterized by low WMH contrast. The ground truth segmentations, initially derived from two existing WMH segmentation tools, were meticulously refined through a precise and time-intensive process of manual thresholding and/or editing. This dataset was further enriched through a variety of data augmentation and resampling techniques. The creation of this large, diverse, and highly accurate ground truth dataset was an important component of the segcsvd_WMH_ development strategy aimed at enhancing segmentation performance.

### Superior agreement with ground truth

The performance of segcsvd_WMH_ and the benchmark tools was evaluated using a comprehensive set of performance metrics, each designed to measure a different aspect of segmentation accuracy. Across a diverse set of test datasets, segcsvd_WMH_ consistently outperformed the benchmark tools, demonstrating significant improvements in each metric. For the 3D isotropic datasets with reduced WMH contrast, these improvements were particularly substantial, with performance metrics for segcsvd_WMH_ exceeding the benchmark tools by 8% to 45%.

Compared to the benchmark tools, segcsvd_WMH_ also displayed stronger and more consistent correlations with periventricular, deep, and total WMH ground truth ROI volumes. In contrast, the benchmark tools showed varying levels of performance across ROIs, with weaker correlations, and/ or biased correlations characterized by a tendency toward underestimation of the true volumes. Furthermore, for HyperMapp3r, strong performance for total and periventricular ROIs was observed alongside much weaker performance for deep WMH ROIs, indicating that global performance measures can obscure important differences in regional sensitivity unless specifically evaluated.

Importantly, segcsvd_WMH_ also exhibited greater agreement with WMH volumes across low, moderate and high levels of total WMH burden compared to the benchmark tools. Altogether, these results suggest that segcsvd_WMH_ may provide more accurate and reliable segmentation performance in complex clinical datasets characterized by varying degrees of CSVD severity.

### Consistent performance across binary segmentation thresholds and datasets

segcsvd_WMH_ exhibited highly robust performance across a wide range of binary segmentation thresholds, maintaining strong and consistent performance metrics across four diverse test datasets. Importantly, for any particular binary segmentation threshold, a similar level of performance was observed for each metric across all datasets. This indicates that segcsvd_WMH_ provides a high degree of stability in segmentation outcomes, potentially eliminating the need for dataset-specific adjustments to the binary segmentation threshold to achieve similar levels of performance across datasets, as can be required for other segmentation tools.

### Robustness to moderate levels of simulated MR artifact

segcsvd_WMH_ also displayed robust performance across a range of mild to moderately severe levels of simulated spike noise artifact, effectively handling data most likely to pass standard quantitative or qualitative quality control procedures. At higher severity levels, segmentation failures for segcsvd_WMH_ were observed whereas HyperMapp3r displayed more accurate and consistent performance under these extreme conditions. While the translation of performance from simulated to real-world artifacts is complex, this result nevertheless suggests that in instances where data is substantially compromised by artifacts, other WMH segmentation tools may be more suitable than segcsvd_WMH_.

### Orientation independence of segcsvd

The performance of segcsvd_WMH_ remained consistent across both the standard and transformed test datasets, showcasing its robustness to changes in image orientation and inaccuracies in the image orientation metadata. For the latter finding to be useful in practice, FreeSurfer’s SynthSeg tool would also need to demonstrate similar robustness, and this was not assessed because preliminary work indicated that inaccuracies in the image orientation metadata resulted in segmentation failures for this tool. Nevertheless, these findings effectively highlight the orientation independence of the core segcsvd_WMH_ segmentation model and validate the success of the orientation augmentation strategy in ensuring consistent performance for image orientations absent from the (non-augmented) training dataset. These findings suggest that segcsvd_WMH_ did not merely learn to segment WMH based on a single, fixed image orientation, but instead developed a deeper, more generalized representation of spatial relationships critical for WMH segmentation, which may have been one factor contributing to its overall success

### Strong generalization performance

The generalization performance of segcsvd_WMH_ was evaluated on two out-of-sample datasets that were excluded from the model training process (2D_OOS_; n=41 and 3D_OOS_; n=5). segcsvd_WMH_ displayed high agreement with ground truth segmentations across all metrics for both datasets, where performance was comparable to that observed for the in-sample test datasets. Additionally, for the qualitative analysis performed on the scans in the 3D dataset without ground truth labels (n=80), segcsvd_WMH_ consistently ranked as the most accurate compared to the two highest-performing benchmark tools. Altogether, these findings highlight the strong generalization capabilities of segcsvd_WMH_ on two challenging out-of-sample test datasets.

### Innovative approach to WMH segmentation

segcsvd_WMH_ segments WMH on FLAIR images, reducing the variability that is introduced by multi-modal segmentation approaches that utilize other scans/image sequences as additional inputs (e.g. FLAIR+T1), rendering it more accessible and streamlined for both clinical and research applications. segcsvd_WMH_ also employs a hierarchical two-stage segmentation framework, leveraging anatomical context in the first stage to enhance WMH segmentation performance in the second stage. This approach significantly outperformed the single-stage benchmark tools, including WMH-SynthSeg, which also segments the two regions included in the segcsvd_WMH_ regional map concurrently with WMH. Future comparison of these methodologies promises to be informative, potentially revealing inherent advantages and disadvantages of concurrent versus hierarchical WMH segmentation strategies.

### Diversity of the training data

HyperMapp3r was trained using a much smaller range of gamma values for contrast augmentation and included only 2D isotropic FLAIR data with high WMH contrast. This approach led to a significant reduction in sensitivity when applied to 3D isotropic FLAIR data with low WMH contrast, and highlights the importance of including a variety of contrast profiles in the training data, as this can be crucial for adequately framing and defining the segmentation task. Without such diversity, deep neural networks can be highly susceptible to the phenomenon of “shortcut learning” (**Geirhos et al., 2020**). This occurs when the network learns an unintended “shortcut” strategy that is only superficially successful and fails under slightly different circumstances. In the context of WMH segmentation, relying primarily on high WMH contrast data for training may cause the network to prioritize global intensity features, potentially neglecting more nuanced features related to local contrast or spatial context which are vital for accurately differentiating WMH from non-WMH on images with reduced WMH contrast in 3D isotropic datasets.

### Challenge and limitations

Despite its strengths, certain limitations of segcsvd_WMH_ should be noted. While a multi-resolution dataset was used to train segcsvd_WMH_, its ability to adapt to data with substantially different voxel sizes, or data acquired at field strengths above 3T, is unknown. Its performance for stroke populations is also unknown and would likely be adversely affected, given that only small infarcts were present in the training dataset. Furthermore, segmentation failures may occur for data affected by substantial MR artifacts, necessitating quality control measures and possibly restricting its applicability for certain datasets. The generalizability of segcsvd_WMH_, while outperforming the benchmark tools, also requires ongoing assessment, and fine-tuning the segmentation model with additional data may be necessary to maintain or improve its efficacy on unseen data.

A further limitation of this study concerns the selection of a finite number of benchmark tools for comparison. While an effort was made to select the most accessible and appropriate alternatives, this selection inherently limits the scope of the comparison. Nevertheless, segcsvd_WMH_ significantly outperformed three other tools which themselves have previously been found to outperform several other established methods. Furthermore, while recently released tools like LST-AI (Wiltgen et al., 2024) were considered, in preliminary testing they were found to miss critical CSVD-specific WMH, but may yield better results on other datasets, particularly when applied to their targeted clinical populations, such as multiple sclerosis (MS).

### Summary of key contributions

This work introduced segcsvd_WMH_ a novel FLAIR-based WMH segmentation tool optimized for enhanced performance across diverse clinical datasets. The strategy for achieving this enhanced performance was twofold. First, in contrast to other machine learning approaches that employ strategies to compensate for noisy labels (**Nagarajan et al., 2024**), significant resources were dedicated to the creation of a large and diverse training dataset with highly accurate, manually refined, ground truth labels. Second, unlike traditional approaches that incorporate minimally processed data from other image modalities (e.g. T1+FLAIR; Mojiri Forooshani et al., 2022), regional anatomical information from a prior FLAIR-based segmentation was incorporated as an additional input to the model.

This approach proved to be highly effective, with segcsvd_WMH_ demonstrating significant performance improvements over the three benchmark tools across a variety of challenging test datasets. Consequently, segcsvd_WMH_ represents a significant step toward more reliable, precise, and adaptable WMH segmentation, demonstrating that substantial performance gains are achievable through the creation of high-fidelity training labels combined with the strategic design of model inputs.

### Future directions

Future segmentation tools are planned to expand upon the hierarchical segmentation framework established by segcsvd_WMH_. These tools will utilize the output of segcsvd_WMH_ output to improve the segmentation accuracy of other CSVD markers. Additionally, the strong performance of segcsvd_WMH_ will be leveraged to create exceptionally large and diverse ground truth datasets, which will be used to train new segmentation models that operate independently of SynthSeg and provide greater robustness to MR artifacts.

## Methods

### MRI data

A large dataset was assembled from seven cohort studies, consisting of FLAIR and FLAIR-based SynthSeg segmentations, from 733 individual patients. This dataset also included T1 images, but these were not used for model development and were instead used as a reference space to co-register and up-sample the FLAIR data, in keeping with in-house imaging processing practices for cases where the resolution of the T1 was superior to that of the FLAIR. Additionally, the T1 images served as a second input for two of the three benchmark tools, as described in the following sections.

This dataset (n=733) was further augmented by additionally resampling the images to have an isotropic voxel size of 1 mm_3_, for a subset of images that were not already at this resolution (n=214). This resulted in a total of 947 FLAIR images (**Table 1**). This inclusion of resampled data served two main objectives. First, it allowed for a multi-resolution training framework, which has the potential to yield models with enhanced adaptability and generalization performance. Second, the inclusion of images intentionally sampled at two different voxel sizes was intended to emphasize essential features that persist across different resolutions, potentially enhancing feature extraction to produce more robust models.

The augmented dataset (n=947) was partitioned into separate training (n=781), validation (n=12), and test (n=154) datasets. Patient data that were sampled at two different resolutions (n=214) were assigned exclusively to either the training, validation or test datasets, ensuring that data from individual patients was not distributed across the partitioned datasets.

The test dataset was further subdivided into four separate datasets based on two factors: whether scans were from a dataset included (“in-sample”) or excluded (“out-of-sample”) from the training dataset, and whether the scans were acquired with approximately isotropic 1mm resolution (3D) or with 3mm thick slices (2D). This resulted in four separate datasets: 2D_IS_, which includes 2D in-sample scans from the VBH, LIPA, MIT, ONDIS, and CAIN datasets, totaling 18 scans; 2D_OOS_, which includes 2D out-of-sample scans from the ONDOOS dataset, totaling 41 scans; 3D_IS_, which includes 3D in-sample scans from the LD dataset, totaling OOS, which includes 3D out-of-sample scans from 10 scans; and 3D the ADNI dataset, with 5 scans having ground truth segmentation and 80 without.

### Image pre-processing

The FLAIR images were processed according to one of two pipelines. In the first pipeline, the FLAIR images were first skull-stripped using ICVMapp3r (Ntiri et al., 2021), bias field corrected using the N4 algorithm (Tustison et al., 2010), and then the masked, bias-corrected images were segmented using FreeSurfer’s SynthSeg tool (**Billot et al., 2023**). In the second pipeline, the FLAIR images were first segmented using FreeSurfer’s SynthSeg tool, then masked with the SynthSeg output, and finally bias field corrected using the N4 algorithm (n=85; 3D_OOS_ test dataset). The second pipeline was adopted later in the project, after the release of the SynthSeg tool, and its integration into the current approach. The use of this pipeline not only streamlined the workflow, reducing total pre-processing time to under 3 minutes inclusive of time required for FreeSurfer’s SynthSeg tool on a 8-core Intel i7-11700 CPU, but also enhanced the pre-processing diversity of the out-of-sample test data. Lastly, after masking and bias correction of the FLAIR image, all foreground voxels were z-score intensity normalized to have a mean of zero and a standard deviation of one.

### WMH contrast Ratios

Contrast ratios for WMH were used to analyze differences in the visibility of WMH relative to grey matter (GM) and white matter (WM) on the FLAIR images across datasets. These were calculated for each FLAIR image as:

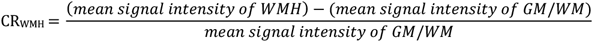

The ground truth segmentations were used to identify WMH voxels and the SynthSeg output was used to identify GM/WM voxels. Larger WMH contrast ratios indicate increased visibility of WMH relative to grey matter (GM) and white matter (WM), suggesting a reduction in the complexity of the segmentation task.

### Model design and training

#### Model architecture

The segmentation model for segcsvd_WMH_ is based on the U-Net architecture with residual units (Falk et al., 2019), which is a convolutional neural net (CNN) designed specifically for medical image segmentation tasks. It is composed of encoder and decoder pathways with skip connections to link the corresponding layers. These skip connections allow the network to capture both local and global features within the input data. The encoder pathway progressively downsamples the input data to extract hierarchical features at multiple spatial scales, while the decoder pathway upsamples the extracted features to generate the final segmentation output. This model, implemented in PyTorch using the MONAI toolkit (**Cardoso et al., 2022**), was configured using: 3 spatial dimensions; 2 input channels; a channel progression of 32, 62, 128, 256, 320; kernel size of 3, strides of 2, 2 residual units, batch normalization and a dropout rate of 0.1. The model takes as input both the pre-processed (masked/bias-corrected) FLAIR image and the SynthSeg segmentation. The SynthSeg segmentation is then masked to include only the sulcal CSF and hippocampal regions. These regions were selected because they provide relevant spatial context but are also sufficiently distanced from the white matter compartment to minimize the potential impact of any errors in the SynthSeg output on the WMH segmentation task.

#### Model training, ensembling, and prediction

Three separate models were trained using the Tversky loss function, which is an extension of the Dice coefficient that measures the similarity between two samples. The Tversky loss (TL) is defined as:

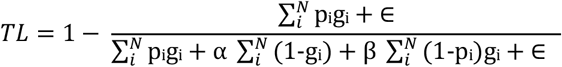

where:

p_i_ and g_i_ are the predicted and ground truth binary values at each pixel i

α and β are parameters that control the relative importance of false positives and false negatives respectively

∈ is a small constant added for numerical stability

N is the total number of pixels

When beta is equal to alpha, the Tversky loss function is equivalent to the Dice coefficient and provides a balanced weighting between false positives and false negatives. When beta is either greater than or less than alpha, more emphasis is placed on minimizing false negatives or false positives, respectively.

The three models were trained using different beta values for the Tversky loss function (β=0.50, 0.55, 0.60), and each model was initialized using a different random seed. This approach was designed to encourage each model to follow slightly different learning trajectories, thereby creating a diverse set of models, each characterized by subtle variations in their trade-off between precision and sensitivity. All other training hyperparameters remained the same across the three models. These hyperparameters included the use of the Adam optimizer with a learning rate of 0.00014, a training duration of 600 epochs, a patch size of 96 voxels^3^, a batch size of 4, a dropout rate of 0.1 and batch normalization. These parameters were empirically chosen based on their efficacy in preliminary experiments using small “toy” datasets, with the rationale that parameters yielding favorable results in this context are likely to generalize well to the full-scale task. This strategy aimed to reduce the risk of overfitting, by selecting parameters with broad applicability rather than finely tuning parameters to a particular validation dataset.

Patch-based training was selected instead of whole-brain training as a strategy to address the issue of class imbalance by maintaining a consistent positive-to-negative label ratio for the WMH class. For each sample in the batch, four patches were extracted using a 3:1 positive-to-negative label ratio.

To enhance model performance in response to variations in image contrast, noise, and orientation, several data augmentation transforms were randomly applied during training, each with a probability of 0.5. These transforms included: contrast adjustment with gamma values randomly sampled between 0.4 and 1.2; the addition of Rician noise with a mean of 1.0 and standard deviation equal to the maximum voxel intensity of the image divided by 10,000; and random 90-degree rotations along each of three principal image axes.

These transforms were selected to simulate the types of variability that are often present in clinical imaging datasets.

After training, a final, composite ensemble model was generated from the three base models. This was accomplished by establishing limits on the number of training epochs that would be considered for model ensembling, to prevent inclusion of models prone to overfitting or underfitting. The lower and upper limits were set to 300 and 525 epochs, respectively. These limits marked the initial performance plateau for the validation data, where the Dice coefficient and sensitivity scores first stabilized, and after which, began to exhibit only marginal improvements. Five models were then randomly selected from within these limits for each base model to form the final composite model (n=15).

To obtain the final segmentation image from the composite model, the input images were first resampled to have an isotropic voxel size of 1 mm^3^. This was implemented as an optional parameter, allowing for flexibility in cases where a alternative voxel sizes would be preferable for specific applications or datasets. Predictions were then generated from each model in the ensemble using a simple test-time augmentation strategy consisting of two different patch sizes: one identical to the patch size used during training (96 voxels^3^), and one larger (128 voxels^3^). This strategy was informed by preliminary experiments which suggested that the use of patches larger than those used during training reduced underestimation in some cases of very high WMH burden. This approach produced 50 predictions per voxel, which were averaged to form a consensus segmentation image. This image was then resampled to match the input resolution, and thresholded at 0.35 to produce the final binary WMH segmentation image. This threshold was selected empirically to provide a balance between thresholds that tended to achieve the highest dice scores (∼0.4) and thresholds that achieved the highest sensitivity scores (∼0.3), on the validation data and in similar segmentation tasks.

On an older generation Intel i7-11700 CPU, segcsvd_WMH_ completes WMH segmentation in approximately 4 minutes, inclusive of the time required for FreeSurfer’s SynthSeg.

### Ground truth generation

For most of the datasets in this work, ground truth WMH segmentations were available. These segmentations were generated by a legacy pipeline (Gibson et al., 2010), and tended to be relatively conservative, often missing smaller lesions or voxels at the edges of WMH. To correct this, one of the benchmark tools, HyperMapp3r, was used in a semi-automated manner to generate an improved set of ground truth WMH segmentations. Skull-stripped, bias-corrected co-registered T1 and FLAIR images were used as inputs to HyperMapp3r, and the HyperMapp3r output was subjected to variable manual thresholding at either 0.3, 0.5, 0.7, or 0.9, to optimize and fine-tune the segmentation result for each individual image. Finally, a consensus image was created that included all voxels present in both the manually thresholded HyperMapp3r output and legacy ground truth image.

For the non-isotropic data (2D_IS_/2D_OOS_), manual thresholding of the HyperMapp3r output improved the capture of true positive voxels, but also introduced occasional false positive voxels. This issue was corrected by removing any 3D connected component on the consensus segmentation image that was not also connected to a WMH voxel on the legacy GT image, with subsequent visual verification of the output.

For the isotropic or nearly isotropic data (3D_IS_/3D_OOS_), only HyperMapp3r segmentations were available. Optimal manual thresholding of the HyperMapp3r segmentation resulted in a considerable number of both false positive and false negative voxels for these datasets, and the correction of these errors required extensive manual editing. This editing was performed sequentially in all three image planes (axial/coronal/sagittal) to ensure the accurate delineation of each WMH as a cohesive 3D object. E.G. performed the initial 3D manual editing, and an experienced image analyst (L.A.W.) reviewed the output and corrected any remaining errors. This was a time-consuming process, requiring between 4 to 16 hours per scan, depending upon WMH burden, and was performed for all 70 scans in the 3D_IS_ dataset (used for both model training and testing), and five scans in 3D_OOS_ dataset (used only for model testing).

### Model performance evaluation

#### Benchmark tools

Three existing WMH segmentation tools, HyperMapp3r, SAMSEG and WMH-SynthSeg, were used to benchmark the performance of segcsvd_WMH_. These tools were selected because they are widely accessible and were all developed specifically with the aim of providing robust performance on diverse clinical datasets without retraining. HyperMapp3r is an earlier iteration of segcsvd_WMH_, based on a similar network architecture and trained on a subset of the current data. SAMSEG is part of the FreeSurfer neuroimaging analysis package and performs simultaneous segmentation of WMH and other cortical and subcortical structures. WMH-SynthSeg was recently added to the development version of the FreeSurfer package (as of December 2023) and is an expanded version of SynthSeg that also performs WMH segmentation.

HyperMapp3r was developed using multi-site, patient data but was limited to older 2D imaging protocols, while SAMSEG and WMH-SynthSeg were developed using synthetic data and an unsupervised approach specifically designed to adapt to data acquired with different imaging protocols (**Laso et al., 2024; Puonti et al., 2016**). Both T1 and FLAIR images were provided as inputs to HyperMapp3r and SAMSEG. This was required for HyperMapp3r and optional for SAMSEG. For SAMSEG, the T1 was included because prior work indicated that this resulted in better performance than using the FLAIR alone (**Puonti et al., 2016**). In addition, the optional lesion-mask-pattern parameter was used for SAMSEG. This was set to “0 1”, indicating that candidate WMH voxels should be considered only if their intensity is brighter than cortical gray matter. This option was strongly recommended in the documentation, and a visual assessment of the results, both with and without this parameter, suggested that it was beneficial for minimizing false positive WMH voxels within cortical gray matter. WMH-SynthSeg requires a single input image. In this work, the FLAIR image was used as the single input, guided by previous findings (Billot et al., 2023) and preliminary tests, which indicated that WMH-SynthSeg produces more accurate segmentation results with FLAIR images as compared to T1 images.

#### Binary segmentation thresholding

All four tools apply a threshold to model probabilities to produce the final binary WMH segmentation output. For segcsvd_WMH_ and HyperMapp3r, this threshold was set to 0.35, which was determined empircally, as described above. For SAMSEG and WMH-Synthseg, their default thresholds of 0.3 and 0.5 respectively were used. WMH-SynthSeg was excluded from the secondary analysis with the lower segmentation threshold because it lacks a user-defined option for threshold adjustment, and initial visual assessments of its segmentation output revealed poor performance that was unlikely to be improved through threshold adjustments.

One further analysis was performed for segcsvd_WMH_ and HyperMapp3r to assess the stability of their segmentation performance across 20 different binary thresholds. The segmentation thresholds were varied systematically in increments of 0.05, between 0 to 0.95. SAMSEG and WMH-SynthSeg were excluded from this analysis, as these tools do not generate WMH probability maps by default, and the extensive time required to generate probability maps for these tools post hoc rendered their inclusion in this analysis unfeasible.

#### Agreement with ground truth segmentations and volumes

The performance of each tool was assessed for each test dataset using fives metrics that measured the agreement with the ground truth segmentations for each of the test datasets. These metrics were selected to provide a comprehensive measure of segmentation performance, in accordance with established recommendations for pixel-level segmentation tasks (**Puonti et al., 2016; Billot et al., 2023**):

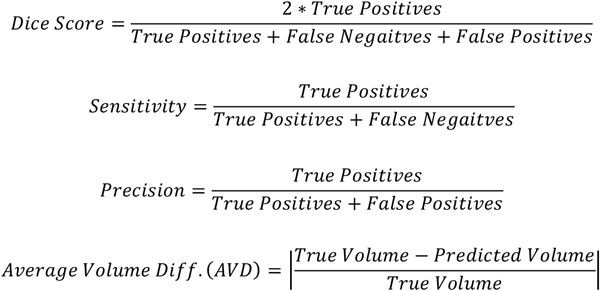

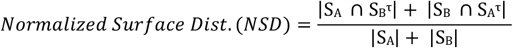

where:

S_A_ = boundary of A

A_B_ = boundary of B

B_B_ ^τ^= border regions of A

B_B_ ^τ^ = border regions of B

S_A_ ⋂ B_B_^τ^ = boundary outside of S_A_ ⋂ B_B_^τ^

S_B_ ⋂ B_A_^τ^ = boundary outside of S_B_ ⋂ B_A_^τ^

τ= 1

The performance of each tool on the full test dataset (n=74) was further assessed by examining the relationship with ground truth volumes for several ROIs that included either periventricular, deep, or total WMH voxels. Periventricular WMH voxels were identified as WM voxels within 1, 2 or 3 voxels from a ventricular CSF voxel, and deep WMH voxels were identified as WMH voxels within 1, 2 or 3 voxels from a cortical GM voxel. This allowed for an assessment of the accuracy of the derived WMH volumes across a total of 7 ROIs.

Given the time-intensive nature of manual tracing for 3D FLAIR data, ground truth data was available for only a small number (n=5) of scans in the ADNI/3D_OOS_ dataset. Consequently, the following two additional analyses were performed to further evaluate segmentation performance in this dataset.

### Qualitative performance evaluation

A trained image analyst (L.A.W.) conducted a detailed qualitative evaluation of the segmentation output for all scans in the ADNI/3D_OOS_ dataset without ground truth labels (n=80). The three top-performing tools, identified by their Dice scores on the standard test data, were included in this analysis. The segmentation output for these tools was anonymized with random filenames, visualized, and subsequently ranked by the analyst in order of most to least accurate.

### Robustness to simulated artifact

The robustness of segmentation performance to increasing levels of data corruption resulting from the addition of simulated spike noise artifacts was examined for the five subjects in the ADNI/3D_OOS_ dataset with available ground truth segmentations. The TorchIO “random spike” data augmentation transform (***Pérez-García et al., 2021***) was used to add simulated spike noise artifacts to the acquired image data. Five separate random spike transforms were generated for each subject by randomly varying two of the transform parameters: the number of spikes (randomly set to either one or two), and the intensity of these spikes (randomly sampled between 1.5 and 2.5). This resulted in five unique transforms that spanned a range of severity levels. These transforms were then applied to the image data (T1/FLAIR/FLAIR-SynthSeg) for each of the five subjects, resulting in a larger, augmented 3D_OOS_ test dataset, each with ground truth segmentations and varying levels of simulated artifact (n=30).

Artifact severity was quantified for the transformed data using the Mattes mutual information (MMI) metric, as implemented by ITK (***Mattes et al., 2003***). In this implementation, a lower MMI value indicates greater similarity between two images. Computing the MMI between the two FLAIR images, with and without simulated artifact, therefore provides a quantitative measure of artifact severity, where smaller MMI values indicate greater similarity, or lower artifact severity, than larger values. Given that the MMI between an image and itself is not guaranteed to be zero, the MMI between the uncorrupted FLAIR image and itself was calculated first, and then subtracted from the MMI between the uncorrupted FLAIR image and each artifact-corrupted FLAIR image. This subtraction resulted in a normalized measure of artifact severity (NMMI) for each transform, where 0 indicates the absence of simulated artifacts, and larger values indicate greater severity of simulated artifacts (**Figure 4)**. Lastly, the NMMI values for each subject were sorted into six levels of artifact severity, from least severe (level 0; no artifacts), to most severe (level 5; max NMMI).

Performance metrics were computed at each artifact severity level and used to assess the robustness of the segmentation performance of each tool, following the methodology introduced in previous work benchmarking the robustness of deep learning models on artifact-corrupted data (***Boone et al., 2023***). Spike noise artifact was selected for this analysis as this transform was not employed during the training phase as a data augmentation strategy for either segcsvd_WMH_ or the benchmark tools. Consequently, this transform provided a challenging “out-of-distribution” test of the robustness of each tool against a type of data corruption not encountered during training.

### Statistical analyses

Permutation tests, consisting of 10,000 permutations, were performed to detect significant differences in the contrast ratios across datasets, and also in the performance metrics for each of the segmentation tools. The criterion for determining statistical significance was defined as a permuted *p*-value less than 0.05. To provide a more precise characterization of statistical significance, permuted *p*-values were reported as *p* < 0.05 or *p* < 0.001.

## Data Availability

This study used data from the Alzheimer's Disease Neuroimaging Initiative (ADNI) database, which is publicly available upon application and approval at adni.loni.usc.edu. Additionally, it incorporated non-public datasets restricted by privacy agreements. Access to these datasets can be requested from the corresponding author under appropriate conditions.

## Author contributions

1) Conceptualization, 2) data curation, 3) formal analysis, 4) funding acquisition, 5) investigation, 6) methodology, 7) project administration, 8) resources & data acquisition, 9) software, 10) supervision, 11) validation, 12) visualization, 13) writing – original draft, 14) writing – review and editing

## Data and code availability statement

A portion of the data used in this work is available from the Alzheimer’s Disease Neuroimaging Initiative (ADNI) database, which can be accessed through an application process on the project website. All other data originates from studies that are not presently publicly available. The code, models, and detailed usage instructions for segcsvd_WMH_ are openly available on GitHub, accessible via the following link: https://github.com/AICONSlab/segcsvd.

## References

Billot B, Magdamo C, Cheng Y, Arnold SE, Das S, Iglesias JE. 2023. Robust machine learning segmentation for large-scale analysis of heterogeneous clinical brain MRI datasets. Proceedings of the National Academy of Sciences 120:e2216399120. doi:10.1073/pnas.2216399120

Boone L, Biparva M, Mojiri Forooshani P, Ramirez J, Masellis M, Bartha R, Symons S, Strother S, Black SE, Heyn C, Martel AL, Swartz RH, Goubran M. 2023. ROOD-MRI: Benchmarking the robustness of deep learning segmentation models to out-of-distribution and corrupted data in MRI. Neuroimage 278:120289. doi:10.1016/j.neuroimage.2023.120289

Cardoso MJ, Li W, Brown R, Ma N, Kerfoot E, Wang Y, Murrey B, Myronenko A, Zhao C, Yang D, Nath V, He Y, Xu Z, Hatamizadeh A, Myronenko A, Zhu W, Liu Y, Zheng M, Tang Y, Yang I, Zephyr M, Hashemian B, Alle S, Darestani MZ, Budd C, Modat M, Vercauteren T, Wang G, Li Y, Hu Y, Fu Y, Gorman B, Johnson H, Genereaux B, Erdal BS, Gupta V, Diaz-Pinto A, Dourson A, Maier-Hein L, Jaeger PF, Baumgartner M, Kalpathy-Cramer J, Flores M, Kirby J, Cooper LAD, Roth HR, Xu D, Bericat D, Floca R, Zhou SK, Shuaib H, Farahani K, Maier-Hein KH, Aylward S, Dogra P, Ourselin S, Feng A. 2022. MONAI: An open-source framework for deep learning in healthcare. doi:10.48550/arXiv.2211.02701

De Guio F, Jouvent E, Biessels GJ, Black SE, Brayne C, Chen C, Cordonnier C, De Leeuw FE, Dichgans M, Doubal F, Duering M, Dufouil C, Duzel E, Fazekas F, Hachinski V, Ikram MA, Linn J, Matthews PM, Mazoyer B, Mok V, Norrving B, O’Brien JT, Pantoni L, Ropele S, Sachdev P, Schmidt R, Seshadri S, Smith EE, Sposato LA, Stephan B, Swartz RH, Tzourio C, van Buchem M, van der Lugt A, van Oostenbrugge R, Vernooij MW, Viswanathan A, Werring D, Wollenweber F, Wardlaw JM, Chabriat H. 2016. Reproducibility and variability of quantitative magnetic resonance imaging markers in cerebral small vessel disease. Journal of Cerebral Blood Flow & Metabolism 36:1319–1337. doi:10.1177/0271678X16647396

Derry A, Krzywinski M, Altman N. 2023. Convolutional neural networks. Nature Methods 20:1269–1270. doi:10.1038/s41592-023-01973-1

Falk T, Mai D, Bensch R, Çiçek Ö, Abdulkadir A, Marrakchi Y, Böhm A, Deubner J, Jäckel Z, Seiwald K, Dovzhenko A, Tietz O, Dal Bosco C, Walsh S, Saltukoglu D, Tay TL, Prinz M, Palme K, Simons M, Diester I, Brox T, Ronneberger O. 2019. U-Net: deep learning for cell counting, detection, and morphometry. Nat Methods 16:67–70. doi:10.1038/s41592-018-0261-2

Fischl B. 2012. FreeSurfer. Neuroimage 62:774–781.

Geirhos R, Jacobsen JH, Michaelis C, Zemel R, Brendel W, Bethge M Wichmann,. 2020. Shortcut learning in deep neural networks. Nature Machine Intelligence 2:665–673. doi:10.1038/s42256-020-00257-z

Gibson E, Gao F, Black SE, Lobaugh NJ. 2010. Automatic segmentation of white matter hyperintensities in the elderly using FLAIR images at 3T. J Magn ResonImaging 31:1311–1322.

Laso P, Cerri S, Sorby-Adams A, Guo J, Mateen F, Goebl P, Wu J, Liu P, Li H, Young SI, Billot B, Puonti O, Sze G, Payabavash S, DeHavenon A, Sheth KN, Rosen MS, Kirsch J, Strisciuglio N, Wolterink JM, Eshaghi A, Barkhof F, Kimberly WT, Iglesias JE. 2024. Quantifying white matter hyperintensity and brain volumes in heterogeneous clinical and low-field portable MRI. doi:10.48550/arXiv.2312.05119

Maier-Hein L, Reinke A, Godau P, Tizabi MD, Buettner F, Christodoulou E, Glocker B, Isensee F, Kleesiek J, Kozubek M, Reyes M, Riegler MA, Wiesenfarth M, Kavur AE, Sudre CH, Baumgartner M, Eisenmann M, Heckmann-Nötzel D, Rädsch T, Acion L, Antonelli M, Arbel T, Bakas S, Benis A, Blaschko MB, Cardoso MJ, Cheplygina V, Cimini BA, Collins GS, Farahani K, Ferrer L, Galdran A, van Ginneken B, Haase R, Hashimoto DA, Hoffman MM, Huisman M, Jannin P, Kahn CE, Kainmueller D, Kainz B, Karargyris A, Karthikesalingam A, Kofler F, Kopp-Schneider A, Kreshuk A, Kurc T, Landman BA, Litjens G, Madani A, Maier-Hein K, Martel AL, Mattson P, Meijering E, Menze B, Moons KGM, Müller H, Nichyporuk B, Nickel F, Petersen J, Rajpoot N, Rieke N, Saez-Rodriguez J, Sánchez CI, Shetty S, van Smeden M, Summers RM, Taha AA, Tiulpin A, Tsaftaris SA, Van Calster B, Varoquaux G, Jäger PF. 2024. Metrics reloaded: recommendations for image analysis validation. Nat Methods 21:195–212. doi:10.1038/s41592-023-02151-z

Mattes D, Haynor DR, Vesselle sH, Lewellen TK, Eubank W. 2003. PET-CT image registration in the chest using freeform deformations. IEEE Trans Med Imaging 22:120–128. doi:10.1109/TMI.2003.809072

Mojiri Forooshani P, Biparva M, Ntiri EE, Ramirez J, Boone L, Holmes MF, Adamo S, Gao F, Ozzoude M, Scott CJM, Dowlatshahi D, Lawrence-Dewar JM, Kwan D, Lang AE, Marcotte K, Leonard C, Rochon E, Heyn C, Bartha R, Strother S, Tardif JC, Symons S, Masellis M, Swartz RH, Moody A, Black SE, Goubran M. 2022. Deep Bayesian networks for uncertainty estimation and adversarial resistance of white matter hyperintensity segmentation. Hum Brain Mapp 43:2089–2108. doi:10.1002/hbm.25784

Nagarajan B, Marques R, Aguilar E, Radeva P. 2024. Bayesian DivideMix++ for Enhanced Learning with Noisy Labels. Neural Networks 172:106122. doi:10.1016/j.neunet.2024.106122

Ntiri EE, Holmes MF, Forooshani PM, Ramirez J, Gao F, Ozzoude M, Adamo S, Scott CJM, Dowlatshahi D, Lawrence-Dewar JM, Kwan D, Lang AE, Symons S, Bartha R, Strother S, Tardif JC, Masellis M, Swartz RH, Moody A, Black SE, Goubran M. 2021. Improved Segmentation of the Intracranial and Ventricular Volumes in Populations with Cerebrovascular Lesions and Atrophy Using 3D CNNs. Neuroinformatics. doi:10.1007/s12021-021-09510-1

Pérez-García F, Sparks R, Ourselin S. 2021. TorchIO: A Python library for efficient loading, preprocessing, augmentation and patch-based sampling of medical images in deep learning. Computer Methods and Programs in Biomedicine 208:106236. doi:10.1016/j.cmpb.2021.106236

Prins ND, Scheltens P. 2015. White matter hyperintensities, cognitive impairment and dementia: an update. Nat Rev Neurol 11:157–165. doi:10.1038/nrneurol.2015.10

Puonti O, Iglesias JE, Van Leemput K. 2016. Fast and sequence-adaptive whole-brain segmentation using parametric Bayesian modeling. Neuroimage 143:235–249. doi:10.1016/j.neuroimage.2016.09.011

Njavants BB, Cook PA, Zheng Y, Egan A, Yushkevich PA, Gee JC. 2010. N4ITK: improved N3 bias correction. IEEE Trans Med Imaging 29:1310–1320. doi:10.1109/TMI.2010.2046908

Chaudhari P, Davatzikos C. 2023. Bias in machine learning models can be significantly mitigated by careful training: Evidence from neuroimaging studies. Proc Natl Acad Sci U S A 120:e2211613120. doi:10.1073/pnas.2211613120

Wardlaw JM, Smith C, Dichgans M. 2019. Small vessel disease: mechanisms and clinical implications. The Lancet Neurology 18:684–696. doi:10.1016/S1474-4422(19)30079-1

Wardlaw JM, Smith EE, Biessels GJ, Cordonnier C, Fazekas F, Frayne R, Lindley RI, O’Brien JT, Barkhof F, Benavente OR, Black SE, Brayne C, Breteler M, Chabriat H, Decarli C, de Leeuw FE, Doubal F, Duering M, Fox NC, Greenberg S, Hachinski V, Kilimann I, Mok V, Oostenbrugge R van, Pantoni L, Speck O, Stephan BCM, Teipel S, Viswanathan A, Werring D, Chen C, Smith C, van Buchem M, Norrving B, Gorelick PB, Dichgans M, STandards for ReportIng Vascular changes on nEuroimaging (STRIVE v1). 2013. Neuroimaging standards for research into small vessel disease and its contribution to ageing and neurodegeneration. Lancet Neurol 12:822–838. doi:10.1016/S1474-4422(13)70124-8

Wiltgen T, McGinnis J, Schlaeger S, Kofler F, Voon C, Berthele A, Bischl D, Grundl L, Will N, Metz M, Schinz D, Sepp D, Prucker P, Schmitz-Koep B, Zimmer C, Menze B, Rueckert D, Hemmer B, Kirschke J, Mühlau M, Wiestler B. 2024. LST-AI: A deep learning ensemble for accurate MS lesion segmentation. NeuroImage: Clinical, 42:103611, doi: 10.1016/j.nicl.2024.103611.

